# High but Short-lived anti-SARS-CoV2 neutralizing, IgM, IgA, and IgG levels among mRNA-vaccinees compared to naturally-infected participants

**DOI:** 10.1101/2022.05.08.22274817

**Authors:** Haissam Abou-Saleh, Bushra Y. Abo-Halawa, Salma Younes, Nadin Younes, Duaa W. Al-Sadeq, Farah M. Shurrab, Na Liu, Hamda Qotba, Nader Al-Dewik, Ahmad H. Ismail, Hadi M. Yassine, Laith J. Abu-Raddad, Gheyath K. Nasrallah

**Affiliations:** Biological Science Program, Department of Biological and Environmental Sciences, College of Arts and Science, Qatar University, Doha P.O. Box 2713, Qatar; Biomedical Research Center, Qatar University, Doha P.O. Box 2713, Qatar; College of Medicine, Q.U. Health, Qatar University, Doha P.O. Box 2713, Qatar; Shenzhen Mindray Bio-Medical Electronics Co., Ltd., Shenzhen, China; Department of Clinical Research, Primary Health Care Centers, Doha, Qatar; Department of Pediatrics, Clinical and Metabolic Genetics, Hamad Medical Corporation, Doha, Qatar; Laboratory Section, Medical Commission Department, Ministry of Public Health, Doha, Qatar; Department of Biomedical Science, College of Health Sciences, QU Health, Qatar University, Doha P.O. Box 2713, Qatar; Infectious Disease Epidemiology Group, Weill Cornell Medicine–Qatar, Cornell University, Qatar Foundation – Education City, Doha, Qatar; World Health Organization Collaborating Centre for Disease Epidemiology Analytics on HIV/AIDS, Sexually Transmitted Infections, and Viral Hepatitis, Weill Cornell Medicine–Qatar, Cornell University, Qatar Foundation – Education City, Doha, Qatar; Department of Healthcare Policy and Research, Weill Cornell Medicine, Cornell University, New York, USA

**Keywords:** mRNA vaccines, waning, neutralizing antibody, Anti-S1-IgA, SRBD-IgM

## Abstract

1.

**Background:** Waning of protection against emerging SARS-CoV-2 variants by pre-existing antibodies elicited due to current vaccination or natural infection is a global concern. Whether this is due to waning of immunity to SARS-COV-2 remains unclear.

**Aim:** We aimed to investigate dynamics of antibody isotype responses among vaccinated naïve (VN) and naturally infected (NI) individuals.

**Methods:** We followed up antibody levels in COVID-19 mRNA-vaccinated subjects without prior infection (VN, n=100) at two phases: phase-I (P-I) at ∼1.4 and phase-II (P-II) at ∼5.3 months. Antibody levels were compared to those of unvaccinated and naturally infected subjects (NI, n=40) at ∼1.7 (P-1) and 5.2 (P-II) months post-infection. Neutralizing antibodies (NTAb), anti-S-RBD-IgG, -IgM, and anti-S-IgA isotypes were measured.

**Results:** VN group produced significantly greater antibody responses (*p*<0.001) than NI group at P-I except for IgM. In VN group, a significant waning in antibody response was observed in all isotypes. There was about ∼ a 4-fold decline in NTAb levels (*p*<0.001), anti-S-RBD-IgG (∼5-folds, *p*<0.001), anti-S-RBD-IgM (∼6-folds, *p*<0.001), and anti-S1-IgA (2-folds, *p*<0.001). In NI group, a significant but less steady decline was notable in NTAb (∼1-folds, *p*<0.001), anti-S-RBD IgG (∼1-fold, *p*=0.005), and S-RBD-IgM (∼2-folds, *p*<0.001). Unlike VN group, NI group mounted a lasting anti-S1-IgA response with no significant decline. Anti-S1-IgA levels which were ∼3 folds higher in VN subjects compared to NI in P-1 (*p*<0.001), dropped to almost same levels, with no significant difference observed between the two groups in P-II.

**Conclusion:** While double dose mRNA vaccination boosted antibody levels, this “boost” was relatively short-lived in vaccinated individuals.

## 2. Introduction

Coronavirus disease 19 (COVID-19) outbreak is the most recent novel infectious disease that placed the world in an emergency state. It is caused by the Severe Acute Respiratory Syndrome Coronavirus-2 (SARS-CoV-2). The disease was declared by the World Health Organization (WHO) as a pandemic on March 11, 2020 (1). Messenger RNA (mRNA) vaccines, such as Pfizer BNT162b2 and Moderna mRNA-1273, are on the top of the list of approved vaccines by WHO and the US Federal Food and Drug Administration (FDA) (2).

On December 21, 2020, Qatar initiated a mass coronavirus disease 2019 (COVID-19) vaccination program, first utilizing the BNT162b2 vaccine and five months later and then the mRNA-1273 (Moderna) vaccine (3-6). Both vaccinations were administered according to the FDA protocol, and vaccine coverage increased progressively from December 2020 until the time of writing (3, 5). Vaccination was rolled out in stages, with frontline health care providers, those with severe or multiple chronic diseases, and people over the age of 70 receiving priority. Vaccination was subsequently expanded to include one age group at a time, as well as certain professional groups (such as teachers), with age serving as the primary criteria for vaccination eligibility throughout the rollout. As of February 26, 2022, it was reported that at least 88% of people aged 12 and over had been fully vaccinated (7).

From January to June 2021, the country had two back-to-back waves that were dominated by the B.1.1.7 (or alpha) and B.1.351 (or beta) variants when vaccination was ramped up (4, 6, 8-11). The B.1.617.2 (or delta) variation was first spotted in the community around the end of March 2021, and by the summer of 2021, it had become the prevalent variety (9-11).

According to current evidence, those who have been fully vaccinated and those previously infected with SARS-CoV-2, have a low risk of infection for at least 6 months before antibody levels start to wane (12). Moreover, recent research has suggested that natural infections protect against reinfection for at least 8–12 months and that vaccination provides substantial protection from the Delta variant (13). However, despite widespread vaccination, the number of people infected with SARS-CoV-2 increased gradually from November 2021 in Qatar, with a significant ten-fold increase in the number of new COVID-19 cases in just two weeks, starting December 29, 2021, till January 12, 2022, in which Qatar hit an all-time high number of new daily cases (14). Therefore, protection against the new variants with pre-existing antibodies from natural infection or vaccination, has become a huge concern.

Although vaccine-induced immunity is being extensively studied, the body of evidence for infection-induced immunity is very limited and is insufficient to establish an antibody titer threshold that shows whether a person is protected from infection (12). In addition, there is currently no FDA-approved serology test that clinicians or the general public may perform to establish if a person is protected from infection (12). This study aimed to investigate the dynamics of antibody response among mRNA-vaccinated naïve (VN) and naturally infected (NI) individuals in Qatar.

## 3. Materials and Methods

### 3.1 Ethical approval and sample collection

The study was reviewed and approved by the Institutional Review Board at Qatar University (QU-IRB 1537-FBA/21). The study included vaccinated naïve (VN) participants (n=100) who had received two doses of the two approved mRNA vaccines in Qatar: BNT162b2 and mRNA-1273. Levels were compared to those of unvaccinated but naturally infected (NI) subjects (n=40). Venous blood samples were collected from vaccines in two phases: phase 1 (P-I) and phase (P-II). For the VN group, P-I samples were collected at 1.4 months (median=6 weeks) and P-II samples were collected at 5.3 months (median=23 weeks) after the second dose. For the NI group, P-I samples were collected at ∼ 1.7 months (median=7 weeks), and P-II samples were collected at 5.2 months (median=22 weeks) post-infection with SARS-CoV-2.

Samples were centrifuged, and the plasma was separated from the whole blood and stored at -80° C until performing the tests. Informed consents were signed by participants before sample collection, and each participant filled out a questionnaire about his/her demographic information in addition to questions related to any previous illness, including COVID-19 infection.

### 3.2 Serology testing

Serological testing was performed using the automated analyzer CL-900i® from Mindray biomedical electronics (15-17) to detect: (1) Neutralizing antibodies (REF. SARS-CoV-2 Neutralizing Antibody 121, Mindray, China) with a WHO conversion factor of 1 AU= 3.31 IU/mL, and the reference range is 10 AU/mL to 400 AU/ml. Phosphate-buffered saline (PBS) was used to dilute the samples that presented readings higher than the mentioned range. (2) Antibodies to the RBD of the S1 subunit of the viral spike protein (IgG (S-RBD)) (catalog No. SARS-CoV-2 Anti-S-RBD IgG122, Mindray, China), with a cut off index for the kit is ≥ 10-1000 AU/ml, and WHO standardization factor of 1.15 BAU/mL. IgA against a recombinant S1 domain of the SARS-CoV-2 was detected using Euroimmun Anti-SARS-CoV-2-ELISA (IgA) (EUR S-IgA) (catalog number: EI 2606–9601 A). The IgA ratio was calculated by dividing the extinction of the sample by the calibrator. Ratios < 8 were considered negative, and ≥ 0.8 were considered positive (18). IgM levels were measured using the automated analyzer Vidas® 3 from BioMeriux Diagnostics (Cat. No. 423833, bioMérieux, France). The results interpretation of Vidas IgM according to test index value (i): i < 1.00 Negative (IgM antibodies to SARS-CoV-2 not detected), i ≥ 1.00 Positive (IgM antibodies to SARS-CoV-2 detected). Architect automated chemiluminescent assay (Abbott Laboratories, USA) was used to test the samples for previous infection by measuring the SARS-CoV-2 anti-nucleoprotein IgG antibodies (anti-N), considering that the IgG antibodies produced against the RBD on the spike protein are different from the IgG antibodies produced against the nucleoprotein of the virus. Therefore, the positive anti-N results of SARS-CoV-2 anti-nucleoprotein IgG antibodies indicate previous exposure to the whole virus (17). Samples with previous infections were excluded from the VN group in this study.

### 3.3 Statistical analysis

GraphPad Prism software (version 9.3.1, GraphPad Software, Inc., San Diego, CA, USA) was used to perform the statistical analysis. Continuous variables were summarized using geometric means and 95% confidence intervals (95% CIs). The collected dataset was subjected to the Shapiro–Wilk normality test in order to evaluate the normality of the data. Due to the absence of normal distribution, nonparametric tests were performed using Wilcoxon rank-sum test for pairwise group comparisons and Mann-Whitney U to test for the differences between independent samples. In the different scatter plots, the central horizontal bar line shows the geometric mean titre, and the error bars show the 95% CIs. All p-values were two-sided, at a significance level of 0.05.

## 4. Results

### 4.1 Participants Characteristics

A total of 140 subjects were included in this study, including 100 VN and 40 NI subjects. VN subjects (n=100) had no previous history of infection (also anti-N negative) and received two doses of either BNT16b2 or mRNA-1273. After receiving the second dose in P-I and P-II, the median weeks were 6 (1.4 months) and 23 (5.3 months) weeks, respectively (Table 1). The VN group consisted of 40% females and 60% males. NI subjects (n=40) were unvaccinated COVID-19-recovered individuals. After receiving the second dose in P-I 1 and P-II, the median weeks were 7 (∼ 1.7 months) and 22 (5.2 months) weeks, respectively. The NI group consisted of 20% females and 80% males (Table 1). Among the 40 NI subjects, 45% were symptomatic (18/40), 20% were pauci-symptomatic (8/40), 20% were asymptomatic (8/40), and 15% had unspecified COVID-19 severity (6/40) (Table 1).

**Table 1.**
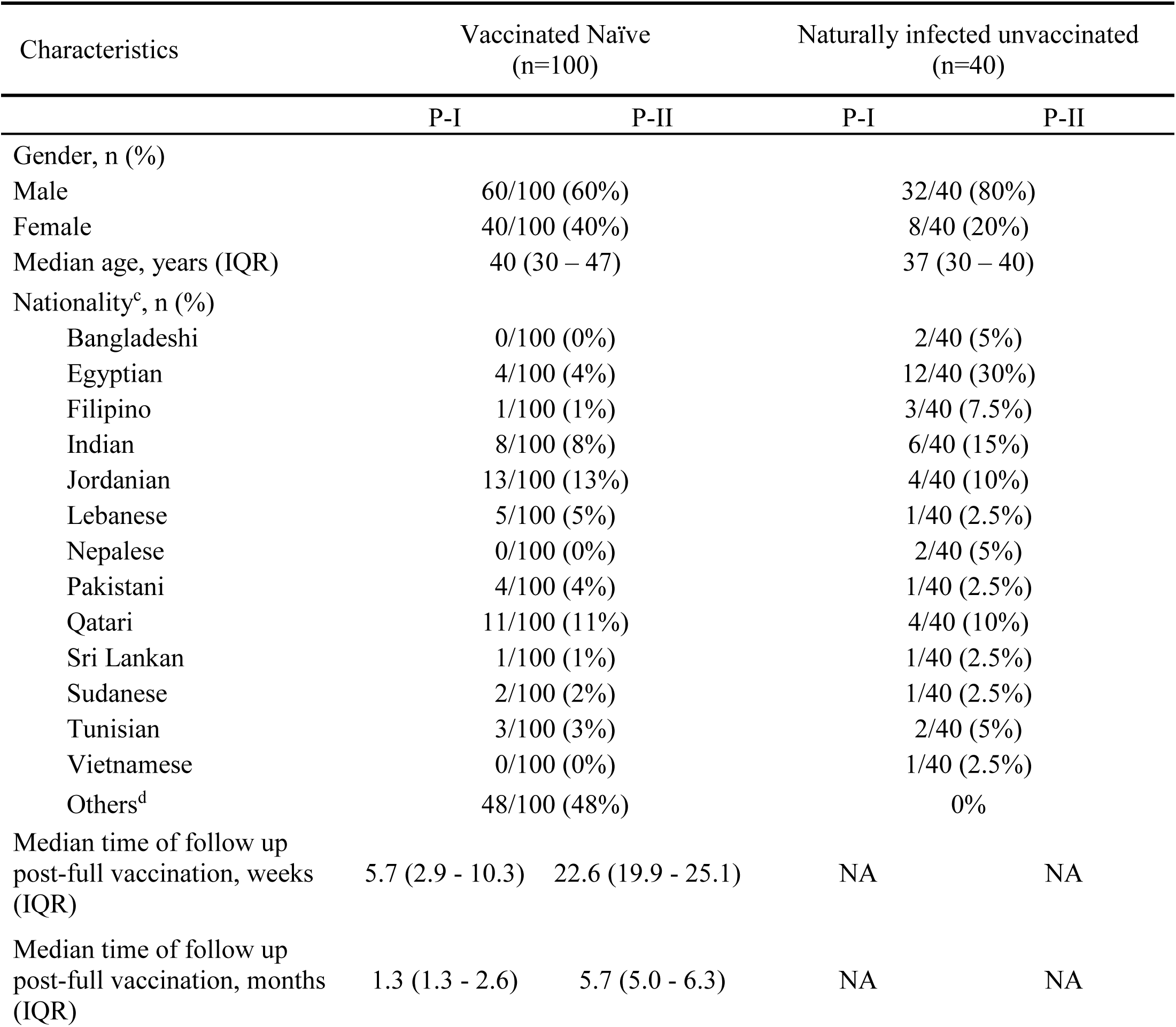
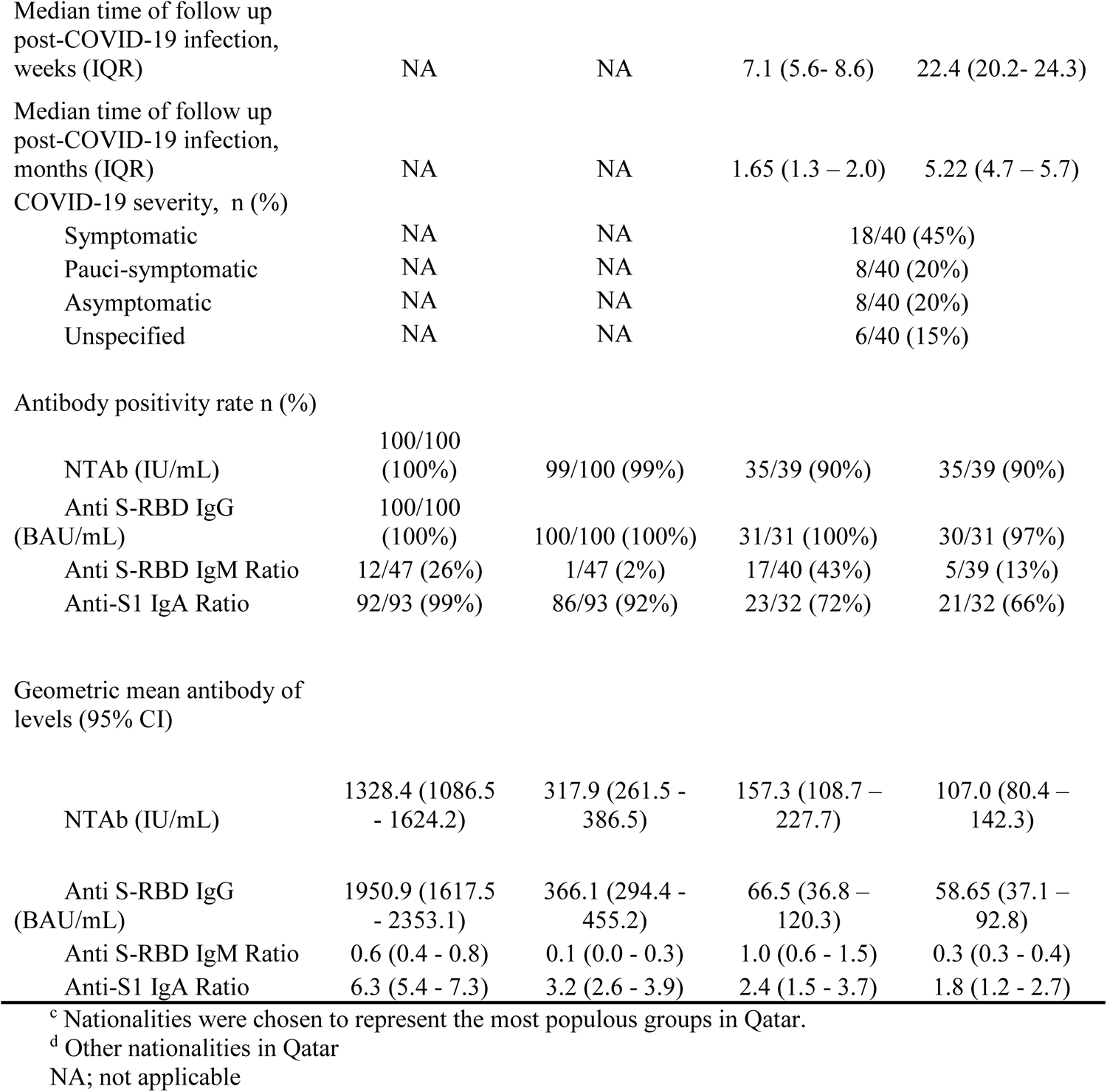
Demographic Characteristics of the study population

### 4.1 Antibody immune response assessment following vaccination

Among the VN participants in P-1 the positivity rates for anti-S-RBD IgG and NTAb antibodies were 100%. For anti-S1-IgA and anti-SRB-IgM, the positivity rates were 99% (92/93) and 26% (12/47), respectively. In P-II, 100% remained positive for anti-S-RBD IgG, and 99% (99/100) were positive for NTAb antibodies. The anti-S1-IgA positivity rate dropped from to 92% (86/93), and only 2% (1/47) remained positive for the IgM.

Among the NI participants in P-1 the positivity rate for NTAb antibodies was 90% (35/39). For anti-S-RBD-IgG, IgM, and IgA, the positivity rates were 100% (31/31), 43% (17/40) and 72% (23/32), respectively. In P-II, 90% (35/39) were positive for NTAb antibodies. The positivity rates dropped to 97% (30/31), 13% (5/39), and 66% (21/32) for anti-S-RBD-IgG, -IgM, and -IgA, respectively.

Among the VN group, waning in antibody levels was significantly observed in all measured parameters (Figure 1). A significant ∼4-fold decrease (*p*<0.001) in the levels of NTAb antibodies was observed, from geometric mean 1328.4 IU/mL (95% CI: 1086.5-1624.2) to 317.94 IU/mL (95% CI: 261.5– 386.5 IU/mL) (Figure 1A). A significant ∼5-fold reduction (*p*<0.001) was observed in the levels of anti-S-RBD-IgG antibody levels from geometric mean 1950.93 BAU/mL (95% CI: 1617.5-2353.1) to 366.107 BAU/mL (95% CI: 294.4– 455.20) (Figure 1B). IgM levels declined by ∼6-fold (*p*<0.001) from geometric mean 0.56 (95% CI: 0.4-0.8) to 0.09 (95% CI: 0.04-0.25) (Figure 1C). The levels of anti-S1-IgA antibodies decreased by ∼2-folds (*p*<0.0001) from geometric mean 6.30 (95% CI: 5.4 – 7.3) to 3.19 (95% CI: 2.6 – 3.9) (Figure 1D).

**Figure 1.**
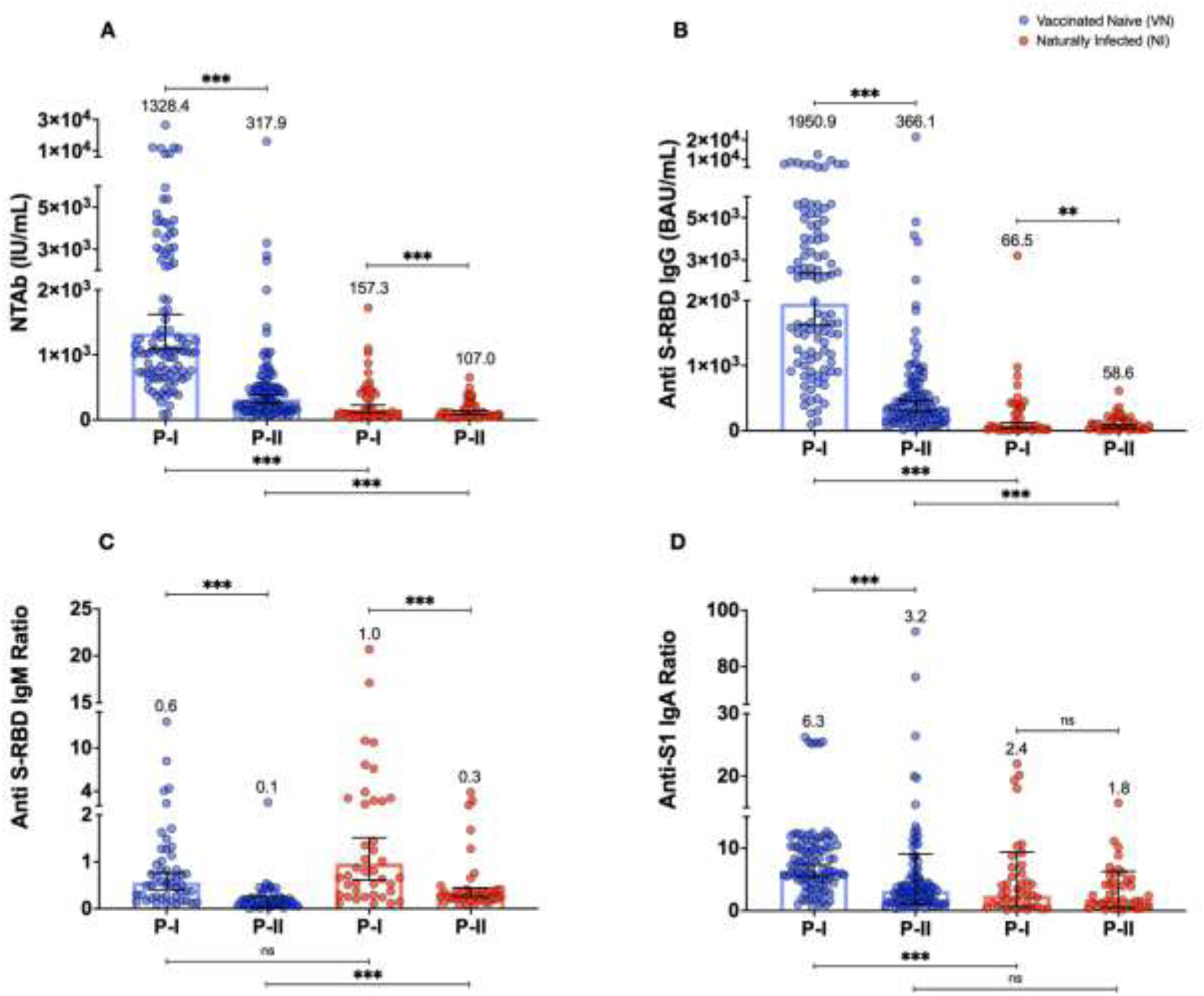
Antibody response measured parameters in P-1 and PII 2 in VN and NI group. **(A)** Neutralizing total antibody levels (IU/mL). **(B)** Anti-S-RBD IgG antibody levels (BAU/mL). **(C)** Anti-S-RBD-IgM levels measured by Vidas. **(D)** Anti-S1 IgA ratios measure by Euroimmune ELISA. Plotted values and horizontal bars indicate the geometric mean and 95% confidence intervals. Statistical significance was determined using two-tailed Wilcoxon matched-pairs signed-rank test. P value asterisk donates to * p≤ 0.05, ** p ≤ 0.01, *** p ≤ 0.001.

Among the NI group, waning in antibody levels was significantly observed in all measured parameters (Figure 1). A significant ∼1-fold decrease (*p*<0.001) in the levels of NTAb was observed, from geometric mean 157.3 IU/mL, 95% CI: 108.7-227.7 to 107.0 IU/mL, 95% CI: 80.42-142.3 (Figure 1A). A significant ∼1-fold reduction was observed in the levels of anti-S-RBD-IgG antibody levels from geometric mean 66.54 BAU/mL (95% CI: 36.81 – 120.3) to 58.65 BAU/mL (95% CI: 37.06 – 92.80) (*p*=0.005) (Figure 1B). IgM levels declined by ∼3-fold from geometric mean 0.96 AU/mL (95% CI: 0.6135 - 1.512) to 0.33 AU/mL (95% CI: 0.2527 - 0.4431) (*p* <0.001) (Figure 1C). No significant decline was observed in anti-S1-IgA levels (Figure 1D).

## 5. Discussion

To the best of our knowledge, this is the first study to comprehensively evaluate the levels of SARS-CoV-2 neutralizing, anti-S-RBD-IgG, Anti-S-RBD-IgM, and anti-S1-IgA antibodies in vaccinated naïve and unvaccinated naturally infected individuals. In the current study, mRNA vaccination elicited significantly greater NTAb, anti-S-RBD-IgG, and anti-S1-IgA levels, compared to natural immunity (Figure 1). These results are in concordance with previous research findings reporting that mRNA vaccines induce higher antibody levels and greater antibody breadth than natural exposure to infection, and differences were particularly notable against the RBD domain (19). In addition, equally remarkable clinical trial data showed rapid induction of mRNA protective efficacy on a timescale similar to our current study (20, 21).

However, despite the mRNA vaccination-boosted antibody levels, this “boost” was relatively short-lived, with ∼2- to 6-fold significant waning in observed in NTAb, anti-S-RBD-IgG, anti-S-RBD-IgM, and anti-S1-IgA levels, 23 weeks post-full-vaccination (Figure 1). These results are in concordance with previous reports of a decline in humoral immune response 3–6 months post-full-vaccination (22-24). IgG and NTAb were reported to significantly decline 3 months post-vaccination by 7- and 4-folds, respectively (22, 23). BNT162b2 and mRNA-1273 vaccines have been reported to elicit potent IgA NTAb in serum after receiving two doses (25, 26). However, consistent with our findings, it was reported that IgA levels declined over time (27).

In comparison to mRNA vaccine-induced immunity, natural infection elicited a significant but less steady ∼1-2-fold decline in NTAb, anti-S-RBD-IgG, and anti-S-RBD-IgM (Figure 1). Interestingly, however, unlike vaccination-induced immunity, a lasting IgA response was observed among naturally infected individuals, with no significant decline observed after ∼22 weeks post-SARS-CoV-2 infection (Figure 1). While anti-S1-IgA levels were ∼3 folds higher in VN subjects compared to NI in P-I (<.001), anti-S1-IgA levels dropped to almost the same between the two groups, with no significant difference observed by the end of the follow-up period.

These findings are similar to a recent report showing that natural infection exhibit a lasting IgA response (28). Furthermore, in natural infection, anti-S1-IgA is known to dominate the early NTAb response (29). Most importantly, it is believed that IgA antibodies have the ability to protect unvaccinated subjects against COVID-19 (30). Recently a study, which examined the patterns of humoral and cellular responses to SARS-CoV-2 for 6 months during the COVID-19 pandemic, reported that subjects who had IgA antibodies only, never succumbed to COVID-19, as opposed to subjects with both IgG antibodies and T cells who contracted the disease (30). It is worth noting that IgA antibody responses in nasal secretions of volunteers infected with the common cold coronavirus 229E have been linked to shorter durations of viral shedding (31). Serum and salivary IgA antibody responses to SARS-Cov-2 spike antigens have recently been reported (32), with salivary IgA antibodies lasting at least three months. Therefore, assessing the immune response of circulating anti-S1-IgA antibodies after vaccination is as important as testing for IgG and IgM.

This study had several limitations. First of all, it is noteworthy to mention that a variety of variables could influence the level of immune response elicited after infection. It should be noted that our NI group included only 45% symptomatic subjects, while the remaining were paucisymptomatic (20%), asymptomatic (20%), or with unspecified severity (15%), which could have affected our results. In those with more severe COVID-19, binding and NTAb antibody titers have been reported to rise faster and reach a greater peak [9, 10, 14]. Individuals with symptomatic SARS-CoV-2 infection have greater antibody titers than asymptomatic, and people who are hospitalized have higher antibody titers than people who are managed as outpatients [9, 10, 15, 16]. Furthermore, several studies have shown a link between cycle threshold (Ct) and antibody titer, with lower Ct values linked with greater antibody titers at the population level [9, 13]. These factors could have impacted the elicited immune response. Furthermore, we have not measured antibody levels prior to vaccine administration.

Despite these limitations, this study has several strengths that merit attention. First, most of the published studies have mainly focused on IgG and IgM, whereas studies on NTAb antibodies, and IgA response are very limited, particularly among naturally infected unvaccinated subjects. Second, in this study, we assessed anti-N antibodies, which is crucial to identify those who had been exposed to a virus but were asymptomatic prior to vaccination, especially among those vaccinated with vaccines containing only S protein. In addition, despite the relatively small sample size across the analyzed groups, we utilized strict inclusion criteria and we included participants from wide age range to achieve valid comparisons.

## 6. Conclusion

Our findings provide important insights into the durability of vaccine- and natural infection-induced immunity. We evaluated the antibody responses of NTAb, anti-SRBD IgM, anti-S1-IgA, and anti-SRBD IgG antibodies. While double-dose mRNA vaccination elicited higher antibody titers compared to natural infection, this “boost” was relatively short-lived in vaccinated individuals. In contrast, natural infection exhibited a less steady decline in NTAb antibodies, IgG, IgM, and a lasting IgA response. Understanding the degree of waning immunity is crucial for policymaking, particularly regarding vaccination strategies, supporting the consideration of booster doses to sustain protection against COVID-19.

## Data Availability

All data produced in the present study are available upon reasonable request to the authors

## Acknowledgment

We thank Ms. Salma Amira Elsharafi, Ms. Fatima AlHamaydeh, Ms. Tala Jamaleddin, and Ms. Huda Abdul Hameed for their help in blood sample collection. We would like also to thank the Biomedical students who participated in this UREP project for their technical support: Ms. Hadiya Khalid, Ms. Nasrin Cusman, Ms.Maram Ali, Ms. Hamas Fouda, Ms. Sumaya Adeeb, and Ms. Salma Muhamoud. We would also like to thank Ms. Sahar Aboalmaaly, Ms. Afra Al Farsi, Ms. Reeham Al-Buainain, Ms. Samar Ataelmannan, and Ms. Jiji Paul, the laboratory technologists at the Medical Commission Laboratory Section, Ministry of Public Health, Qatar, for their help in performing the Architect® immunoassay.

